# Regional differences in reported Covid-19 cases show genetic correlations with higher socio-economic status and better health, potentially confounding studies on the genetics of disease susceptibility

**DOI:** 10.1101/2020.04.24.20075333

**Authors:** Abdel Abdellaoui

## Abstract

**Background:** In March 2020, England showed a rapid increase in Covid-19 cases. Susceptibility for infectious diseases like Covid-19 is likely to be partly genetic. Mapping the genetic susceptibility for Covid-19 outcomes may reveal biological mechanisms that could potentially aid in drug or vaccine developments. However, as the disease spreads unevenly across the country, regional allele frequency differences could become spuriously associated with disease prevalence.

**Methods:** A regional genome-wide association study (RGWAS) was conducted in 396,042 individuals from England to investigate the association between 1.2 million genetic variants and regional differences in daily reported Covid-19 cases from March 1^st^ to April 18^th^ 2020.

**Results:** The polygenic signal increases during the first weeks of March, peaking at March 13^th^ with the measured genetic variants explaining ∼3% of the variance, including two genome-wide significant loci. The explained variance starts to drop at the end of March and reaches almost zero on April 18^th^. The majority of this temporary polygenic signal is due to genes associated with higher educational attainment and better health.

**Conclusions:** The temporary positive relationship between Covid-19 cases and regional socio-economic status (SES) at the beginning of the Covid-19 outbreak may reflect *1)* a higher degree of international travelers, *2)* more social contacts, and/or *3)* better testing capacities in higher SES regions. These signals are in the opposite direction of expected disease risk increasing effects, which has the potential to cancel out signals of interest. Genetic association studies should be aware of the timing and location of cases as this can introduce interfering polygenic signals that reflect regional differences in genes associated with behavior.

## INTRODUCTION

Covid-19 is the infectious disease caused by the single-stranded RNA virus severe acute respiratory syndrome coronavirus 2 (SARS-CoV-2).^1^ The virus was first detected in Wuhan, China, in December 2019,^2^ and soon thereafter spread to other parts in the world. The World Health Organization (WHO) declared a Public Health Emergency of International Concern (PHEIC) on January 30^th^ and a worldwide pandemic on March 11^th^ 2020. In England, the first cases were identified in two tourists visiting York on January 29^th^, on January 30^th^ a public health information campaign was launched to advise people on how to slow the spread of the virus, the first transmission was confirmed February 28^th^, and a rapid increase in infections followed in the beginning of March, raising the UK risk level from moderate to high on March 12^th^.^3^ On March 18^th^, schools in the UK were ordered to close, and on March 20^th^ restaurants, pubs, clubs, and indoor sport and leisure facilities also had to close their doors. As of April 18^th^, England had 95,297 reported Covid-19 cases and 15,464 reported Covid-19 related deaths.

There is much variation in the severity of Covid-19 symptoms, ranging from asymptomatic to mild flu-like symptoms to critical illness. The severity of the symptoms and the mortality rate are strongly associated with age, sex, and underlying health problems. It is not yet clear to which extent genetic susceptibility plays a role in the individual differences in Covid-19 symptoms. It is well-established however that the underlying health problems that are associated with Covid-19 symptom severity, such as obesity, diabetes, and cardiovascular disorders, are heritable complex traits that are caused by a combination of many genes with small effects and environmental influences.^4^ Identifying which genes are associated with Covid-19 outcomes is of great importance, as genetics are likely to play a role in individual differences in the susceptibility for infectious diseases in general,^5^ and drug targets with support from genetic association studies are more than twice as likely to succeed.^6,7^ An international effort to investigate the role of genetic susceptibility for Covid-19 is on the way,^8^ with UK Biobank^9^ as one of the contributing cohorts. Individual-level Covid-19 test results from UK Biobank participants are still ongoing and are not part of the current study; instead, here we use the genotype data and geographic location of UK Biobank participants and link those to the total number of regional Covid-19 cases as reported by Public Health England (PHE)^10^. As one of the largest genotyped datasets in the world with a rich collection of phenotypic measurements (N ∼500,000),^9^ UK Biobank is likely to be contributing a substantial part of the polygenic signal for Covid-19 susceptibility. It is therefore important that the possible sources of polygenic signal in datasets like these are well characterized. UK Biobank provides a unique data resource to address questions regarding the geographic distribution of polygenic complex trait variation in ways that have not been possible before. UK Biobank allowed us recently to identify that, after controlling for ancestry, polygenic complex trait variation is not randomly distributed across geographic space in Great Britain.^11^ The strongest regional differences we observed were in line with regional differences in socio-economic status.^11^ Of the 33 complex traits and diseases we analyzed, polygenic signals associated with educational attainment showed the strongest geographic clustering after controlling for regional ancestry differences.^11^ People that migrated out of the poorest regions in Great Britain had the highest polygenic scores for a higher education level on average, while those that stayed had the lowest on average, a process that increased these regional genetic differences over time.^11^ Regional differences in health outcomes like obesity and diabetes were more in line with regional differences in genes associated with educational attainment than with genes associated with the health outcomes themselves, suggesting environmental influences that cause people from regions with lower education levels to have worse health outcomes.^11^ Obesity and diabetes have been associated with worse symptoms and higher fatality rates for Covid-19,^12,13^ raising the questions about whether we can expect regional differences in Covid-19 outcomes to show similar patterns.

This study investigates whether regional differences in daily reported Covid-19 cases in England during March and the first half of April 2020 are in line with regional genetic differences in UK Biobank participants. Since this study uses regional reports of Covid-19 cases instead of individual-level Covid- 19 outcomes, we expect to have more power to detect regional genetic correlates of social stratification if they are in line with Covid-19 outcomes and less power to detect strong polygenic signals directly related to the genetic susceptibility of Covid-19. If regional patterns of social stratification lead to associations between regional Covid-19 outcomes and genetic variants, these should be accounted for when the aim is to identify genes related to Covid-19 susceptibility in order to better understand the disease and accelerate drug target and vaccine developments.

## RESULTS

### Data and Analysis

Regional Genome-Wide Association Studies (RGWASs)^11^ were run on the cumulative number of reported Covid-19 cases for every day between March 1^st^ and April 18^th^ 2020. The RGWASs were run on 396,042 UK Biobank^9^ participants of European descent on 1,246,531 common single-nucleotide polymorphisms (SNPs) with minor allele frequencies (MAF) > .01. In an RGWAS, all subject get assigned the same phenotype as the rest of the subjects in their region. The phenotypes were defined as 1) the regional daily cumulative number of reported Covid-19 cases, and 2) the regional daily cumulative number of reported Covid-19 cases divided by the regional adult population size according to 2018 census data (case-rate). Regional data on the number of cases were available for 151 local authority regions in England obtained from Public Health England (PHE)^10^ and regional data on the population sizes for 149 out of the 151 regions were obtained from the Office of National Statistics (ONS)^14^. All RGWASs were run using the linear-mixed model approach in fastGWA.^15^ To control for confounding due to population stratification and family-relatedness, we corrected for a sparse genetic-relatedness matrix (GRM) and the first 100 principal components (PCs) and applied an LDSC-intercept based genomic control (GC).^11^ We then computed the SNP-based heritability and the genetic correlation with educational attainment (EA), based on the EA3 GWAS^16^ excluding all British cohorts (N=245,612, SNP-based heritability = .10), using LD Score regression^17,18^. For more details on the methods, rationale, and interpretation of the RGWAS approach, see Abdellaoui et al (2019)^11^.

### Longitudinal SNP-based heritability

The upper panel in Figure 1 shows the longitudinal change in SNP-based heritability estimates from the RGWAS signals from March 1^st^ to April 18^th^. On the first day that was analyzed, there were 82 reported Covid-19 cases in England^10^ and a low SNP-based heritability (case-rate: *h*_2_=0.3%, SE=0.11; total cases: *h*_2_=0.4%, SE=0.13). The SNP-based heritability increased in the weeks that followed, and reached its peak on March 12^th^ for the total cases (*h*_2_=1.4%, SE=0.14) and on March 13^th^ for the case-rate (*h*_2_=2.7%, SE=0.18; see Figure 2). The total number of reported cases in England on March 13^th^ was 1,959, with the highest rates in Westminster and the Royal Borough of Kensington and Chelsea (see Figure 2). While cases continued to rise, the SNP-based heritability remained relatively stable for more than a week, and began to drop after March 20^th^, which had 6,819 reported cases. On April 18^th^, the SNP-based heritability dropped down to 0.4% and 0.1% for the case-rate and the total number of cases respectively.

**Figure 1:**
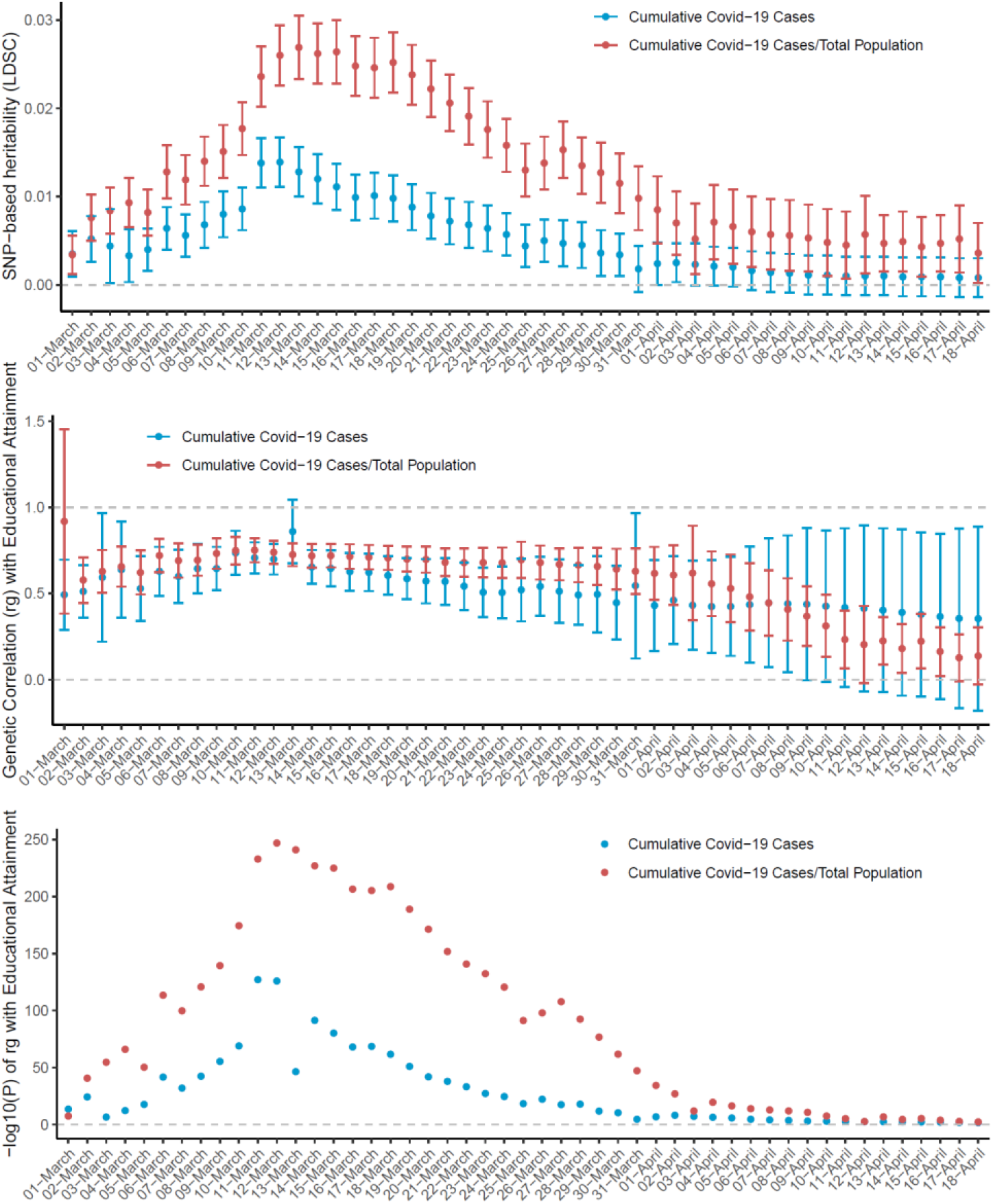
SNP-based heritability of the RGWAS of the daily regional cumulative reported Covid-19 cases and their genetic correlations with educational attainment (EA3^16^ without British cohorts). Error bars are 2×SE.

**Figure 2:**
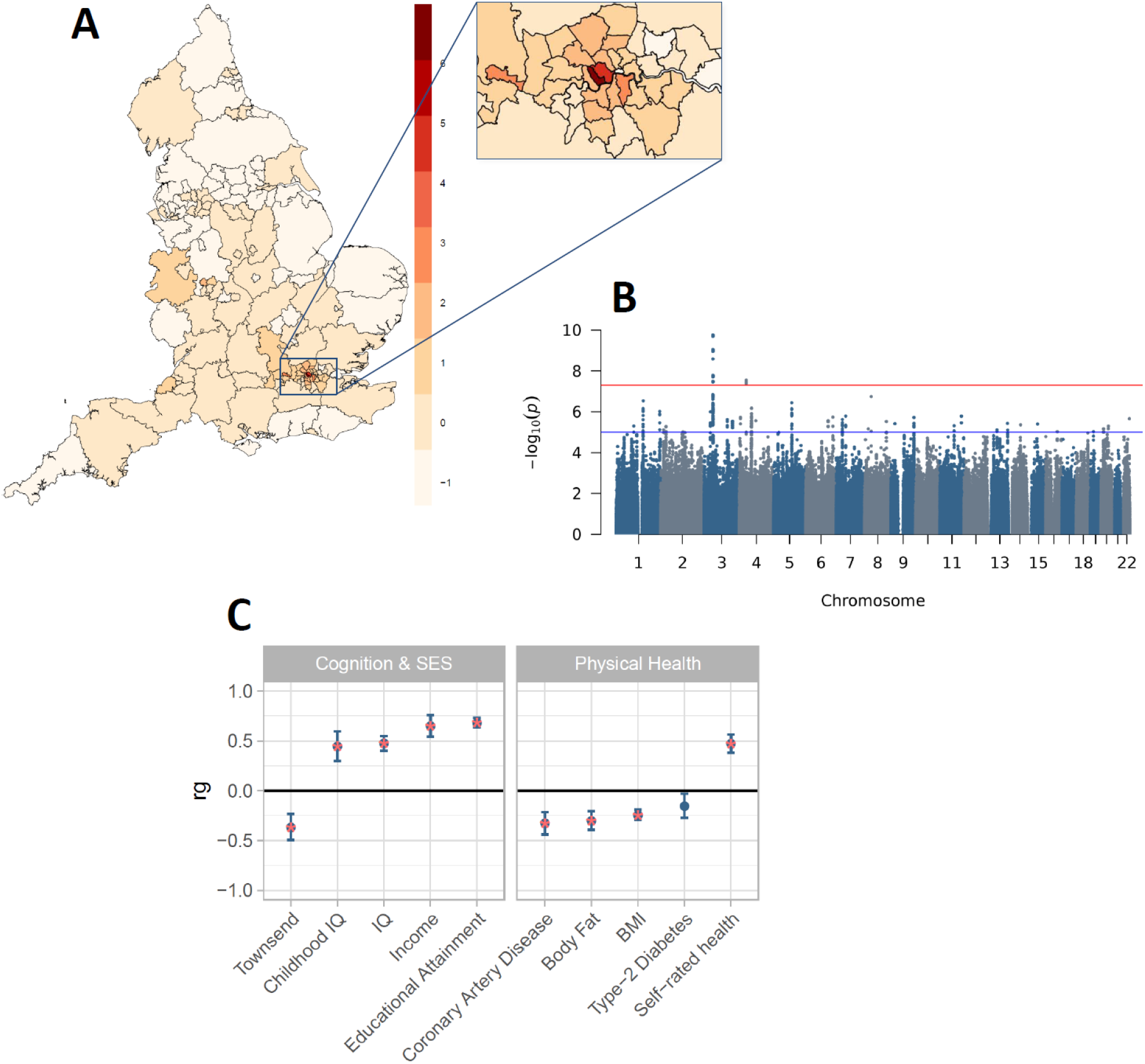
Case-rates on March 13^th^, the day with the highest SNP-based heritability. **A:** Geographic distribution of the reported Covid-19 case-rates on March 13^th^, the day with the highest SNP-based heritability estimate. Color bar indicates distribution frequency of case-rate, scaled such that it has a mean of 0 and SD of 1. **B:** Manhattan Plot of the RGWAS of case-rates on March 13^th^. The suggestive significance threshold (blue line) is set at 1 × 10^−5^, and the genome-wide significance threshold (red line) is set at 5 × 10^−8^. **C:** Genetic correlations (r_g_) of the RGWAS on Covid-19 case-rate on March 13^th^ with 5 SES-related and 5 physical health traits as computed with LDSC regression (red stars indicate significance after Bonferroni correction, i.e., p < .005).

### Genetic correlations

The bottom two panels in Figure 1 show the longitudinal change in the genetic correlation (*r*_g_) between the RGWASs for the regional daily cumulative reported Covid-19 cases and the individual-level educational attainment GWAS (EA3)^16^ without British cohorts. The *r*_g_ with educational attainment showed a similar trajectory as the SNP-based heritability with respect to its significance (bottom panel of Figure 1). The point estimate of the *r*_g_ showed less variability and was .73 on March 13^th^. In April, it started to decrease, reaching .14 on April 18^th^. The standard error increased when the SNP-based heritability decreased, likely due to a weaker genetic signal. The high *r*_g_ with educational attainment is in line with the regional differences in polygenic signals associated with educational attainment that we recently observed for other regional measures of SES, health, and cultural outcomes as well.^11^

For the RGWAS results of March 13^th^, we investigated the genetic correlations with a total of 10 traits, of which 5 traits are related to socio-economic status, and 5 traits are associated with health outcomes that are considered risk factors for worse Covid-19 outcomes (Figure 2C). All genetic correlations were significant after Bonferroni correction, except for Type-2 Diabetes. The genetic correlations were all in the direction that would be expected given a positive association with higher SES^11^, namely higher reported Covid-19 case-rates are in line with higher cognitive abilities, higher income, and better health outcomes (lower BMI, body fat, and cardiovascular risk, and higher self - rated health).

## DISCUSSION

We showed previously that social stratification and selective migration driven by socio-economic status (SES) has likely led to growing regional differences in genome-wide complex trait variation, especially for genes associated with educational attainment.^11^ Results of the current analyses suggest a temporary positive genetic relationship between the reported number of Covid-19 cases in March 2020 and regional SES and health. The strength of this genetic signal increased after controlling for population density by dividing the total number of regional cases by the regional population size. The positive genetic relationship between number of cases and higher SES and health increases in the beginning of the outbreak and then decreases as the number of cases rapidly rises and the disease spreads across the country. These results could reflect that **1)** higher SES regions are more likely to have international travellers, including tourists, **2)** people in higher SES regions are more likely to have higher levels of social contacts, or that **3)** higher SES regions were more likely to have better testing capacities in the beginning of the pandemic. These genetic signals are in the other direction than one would expect based on epidemiological data on risk factors for more severe Covid-19 symptoms,^12,13^ which could supress genetic signals of interest when looking for polygenic signals associated with increased Covid-19 susceptibility.

The results confirm that the large UK Biobank dataset can be leveraged to combine real-time demographic data with genetic data through RGWAS to study the relationship between any regional dynamic variable and social stratification on a national level with high temporal resolution. The RGWAS approach should not be considered as an alternative to the traditional individual-level GWAS, as regional differences are of another nature than individual differences.^11^ Rather, the presence of polygenic signals in RGWAS results should serve as an indicator to proceed cautiously when interpreting the results of individual-level GWASs. The same limitations that were described in the last implementation of the RGWAS approach remain^11^, including the ascertainment bias in the UK Biobank towards a more healthy and higher educated population and the unknown bias in the regional data (in this case, the bias in the regional data may be related to testing capacity and/or hospital access), and gene-environment correlations inflating genetic signals.

This study combines longitudinal regional-level data with individual-level genotype data and GWAS summary statistics from 10 traits related to SES and health to investigate the nature of the polygenic effects associated with reported regional differences in Covid-19 infections. As large individual-level GWASs of Covid-19 susceptibilty are on the way,^8^ it is important to be aware that polygenic signals of Covid-19 susceptibility can contain genome-wide significant signals that reflect social differences in disease prevalence and/or testing capacity, and that these signals can vary over time.

## MATERIALS AND METHODS

### Participants

The participants of this study come from UK Biobank (UKB),^19,20^ which has received ethical approval from the National Health Service North West Centre for Research Ethics Committee (reference: 11/NW/0382). A total of 502,536 participants (273,402 females and 229,134 males) aged between 37 and 73 years old were recruited in the UK between 2006 and 2010. The participants were recruited across 22 assessment centers throughout Great Britain in order to cover a variety of different settings providing socioeconomic and ethnic heterogeneity and urban–rural mix. They underwent a wide range of cognitive, health, and lifestyle assessments, provided blood, urine, and saliva samples, and will have their health followed longitudinally.

### Genotypes and Quality Control (QC)

A total of 488,377 UKB participants had their genome-wide single nucleotide polymorphisms (SNPs) genotyped on either the UK BiLEVE array (N = 49,950) or the UK Biobank Axiom Array (N = 438,423). The genotypes were imputed using the Haplotype Reference Consortium (HRC) panel as a reference set (pre-imputation QC and imputation are described in more detail in Bycroft et al, 2018).^20^ We extracted SNPs from HapMap3 CEU (1,345,801 SNPs) were filtered out of the imputed dataset. We then did a pre-PCA QC on unrelated individuals, filtering out SNPs with MAF < .01 and missingness > .05, leaving 1,252,123 SNPs. After filtering out individuals with non-European ancestry, we repeated the SNP QC on unrelated Europeans (N = 312,927), filtering out SNPs with MAF < .01, missingness >.05 and HWE p < 10^−10^, leaving 1,246,531 SNPs. The HWE p-value threshold of 10^−10^ was based on: http://www.nealelab.is/blog/2019/9/17/genotyped-snps-in-uk-biobank-failing-hardy-weinberg-equilibrium-test. We then created a dataset of 1,246,531 QC-ed SNPs for 456,064 UKB subjects of European ancestry.

### Ancestry & Principal Component Analysis

To capture British ancestry, we first excluded individuals with non-European ancestry. Ancestry was determined using Principal Component Analysis (PCA) in GCTA. The UKB dataset was projected onto the first two principal components (PCs) from the 2,504 participants of the 1000 Genomes Project, using HM3 SNPs with minor allele frequency (MAF) > 0.01 in both datasets. Next, participants from UKB were assigned to one of five super-populations from the 1000 Genomes project: European, African, East-Asian, South-Asian, or Admixed. Assignments for European, African, East-Asian, and South-Asian ancestries were based on > 0.9 posterior-probability of belonging to the 1000 Genomes reference cluster, with the remaining participants classified as Admixed. Posterior-probabilities were calculated under a bivariate Gaussian distribution where this approach generalizes the k-means method to take account of the shape of the reference cluster. We used a uniform prior and calculated the vectors of means and 2×2 variance-covariance matrices for each super-population. A total of 456,064 subjects were identified to have a European ancestry. A PCA was then conducted on the individuals of European ancestry (N = 456,064) in order to capture ancestry differences within the British population (see Figure 3). In order to capture ancestry differences in homogenous populations, genotypes should be pruned for LD and long-range LD regions removed.^21^ The LD pruned (r^2^ < .2) UKB dataset without long-range LD regions consisted of 131,426 genotyped SNPs. The PCA to construct British ancestry-informative PCs was conducted on this SNP set for unrelated individuals using flashPCA v2.^22^ PC SNP loadings were used to project the complete set of European individuals onto the PCs.

**Figure 3:**
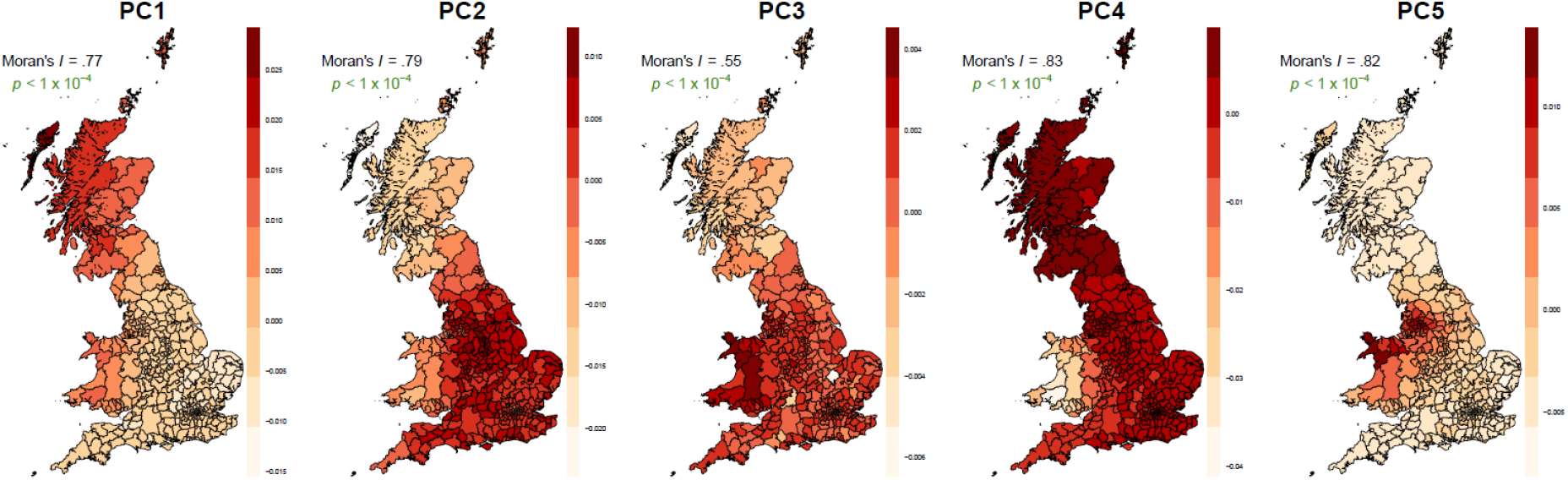
The geographic distributions (birthplace) of the first five PCs, Moran’s I and empirical p-values for Moran’s I. P-values denoted in green are significant after Bonferroni correction (N = 312,927 unrelated individuals of European descent). ^11^

### Genetic Relatedness Matrix

The genetic relatedness matrices (GRMs) contain genetic relationships between all individuals based on a slightly LD pruned HapMap 3 SNP set (LD-pruning parameters used in PLINK: window size = 1000 variant count, step size = 100, r2 = 0.9 and MAF > 0.01, resulting in 575,293 SNPs). The GRMs were computed using GCTA.^23^ We created a sparse GRM, containing only the relationships of related individuals (cut-off = .05, resulting in 179,609 relationships).

### Regional Genome-Wide Association Study (RGWAS)

For the RGWASs, we ran linear mixed model (LMM) GWASs using fastGWA^15^ on participants with European ancestry, which controls for cryptic relatedness and population stratification by including a genetic relatedness matrix (GRM) in the model.^24^ The phenotypes were defined as the number or rate of reported Covid-19 cases in the local authority of the subject’s current address. Sex and age were included as covariates, as were the first 100 PCs as an additional control for population stratification. As observed before with the RGWAS approach,^11^ the results revealed a considerable inflation of test statistics that was not due to polygenic effects, which was captured by LD score intercepts^25^. We controlled for this inflation with an LD score intercept-based genomic control,^25^ i.e., we adjusted the standard errors (SE) of the estimated effect sizes as follows: 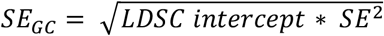. For more details on the methods, rationale, and interpretation of the RGWAS approach, see Abdellaoui et al (2019)^11^.

### SNP-based heritability and genetic correlations

SNP-based heritabilities and genetic correlations were computed using LD-score regression^26^.^26^ The genetic correlation between traits is based on the estimated slope from the regression of the product of z-scores from two GWASs on the LD score and represents the genetic covariation between two traits based on all polygenic effects captured by the included SNPs. The genome-wide LD information used by these methods were based on European populations from the HapMap 3 reference panel. ^25,26^ All LD score regression analyses included the 1,290,028 million genome-wide HapMap SNPs used in the original LD score regression studies.^25,26^ The GWAS summary statistics for the traits for which we ran the genetic correlations (Figure 2C) are from: educational attainment,^16^ excluding all British cohorts (N=245,612, SNP-based *h*_2_=.10), the Townsend^27^ (N=112,151, SNP-based *h*_2_=.04), Childhood IQ^28^ (N=12,441, SNP-based *h*_2_=.28), Adult IQ^29^ (N=78,308, SNP-based *h*_2_=.19), Income^27^ (N=112,151, SNP- based *h*_2_=.06), Coronary Artery Disease^30^ (N=86,995, SNP-based *h*_2_=.28), Body Fat^31^ (N=100,716, SNP- based *h*_2_=.10), BMI^32^ (N=322,154, SNP-based *h*_2_=.13), Type-2 Diabetes^33^ (N=69,033, SNP-based *h*_2_=.18), Self-rated health^34^ (N=111,749, SNP-based *h*_2_=.10).

## Data Availability

UK Biobank data and regional data on Covid-19 cases provided by Public Health England were analyzed, which are both publicly available datasets.

https://coronavirus.data.gov.uk/

## ACKNOWLEDGEMENTS

This study has been conducted using UK Biobank resource under Application Number 40310. UK Biobank was established by the Wellcome Trust medical charity, Medical Research Council, Department of Health, Scottish Government and the Northwest Regional Development Agency. It has also had funding from the Welsh Assembly Government, British Heart Foundation and Diabetes UK. A.A. is supported by the Foundation Volksbond Rotterdam and by ZonMw grant 849200011 from The Netherlands Organisation for Health Research and Development.

